# Sequence Proven Reinfections with SARS-CoV-2 at a Large Academic Center

**DOI:** 10.1101/2022.05.17.22275210

**Authors:** C. Paul Morris, Raghda E. Eldesouki, Amary Fall, David C. Gaston, Julie M. Norton, Nicholas Gallagher, Chun Huai Luo, Omar Abdullah, Eili Y. Klein, Heba H. Mostafa

## Abstract

**Background:** Increased reinfection rates with SARS-CoV-2 have recently been reported, with some locations basing reinfection on a second positive PCR test at least 90 days after initial infection.

**Methods:** We identified cases where patients had two positive tests for SARS-CoV-2 and evaluated which of these had been sequenced as part of our surveillance efforts, and evaluated sequencing and clinical data.

**Results:** 750 patients (920 samples) had a positive test at least 90 days after the initial test. The median time between tests was 377 days, and 724 (79%) of the post 90-day positives were collected after the emergence of the Omicron variant in November 2021. Sequencing was attempted on 231 samples and successful in 127. Successful sequencing spiked during the Omicron surge and showed higher median days from initial infection compared to failed sequences (median 398 days compared to 276 days, p<0.0005). A total of 122 (98%) patients showed evidence of reinfection, 45 of which had sequence proven reinfection and 77 had inferred reinfections (later sequence showed a clade that was not circulating when the patient was initially infected). Children accounted for only 4% of reinfections. 43 (96%) of 45 infections with sequence proven reinfection were caused by the Omicron variant, 41 (91%) were symptomatic, 32 (71%), were vaccinated prior to the second infection, and 6 (13%) were Immunosuppressed. Only 2 (4%) were hospitalized, and both had underlying conditions.

**Conclusion:** Sequence proven reinfections increased with the Omicron variant but generally caused mild infections.

## Introduction

The Omicron variant of SARS-CoV-2 quickly displaced the Delta variant to become the most predominant by the end of December 2021 [1]. The factors that contributed to the unprecedented success of Omicron are not completely understood, but immune evasion is the most likely. Some reports showed a decrease in antibody neutralization to Omicron in both vaccinated and previously infected individuals [2, 3]. Additionally, a large increase in the rates of breakthrough cases and reinfection with the Omicron variant was reported [4, 5]. In most cases of reinfections, sequencing data was not available to validate these findings. Thus, reinfections were historically suspected, generally, by a positive PCR over 90 days after the original PCR in asymptomatic individuals, or 45 days after initial infection in symptomatic individuals [6]. There are potential issues with this approach as prolonged shedding of RNA and prolonged active infection are both well documented [7, 8], and can last longer than 90 days [9]. It is therefore unclear in these cases whether the positive result could be due to prolonged shedding, repeat infection, or erroneous PCR results.

We previously reported infrequent cases of sequence proven reinfections, and a few instances where positivity over 90 days is consistent with persistence of the genome from the initial infection [9] using data obtained prior to the Omicron surge. In this study, we evaluate SARS-CoV-2 genomes of cases of individuals who have tested positive at least 90 days from an initial positive, to determine the frequency of reinfections. We further characterize vaccination status, age, immune status, and outcomes in patients with sequence proven reinfections, along with time from initial infection and dates at which reinfections took place.

## Methods

### Sample Criteria/Inclusion

SARS-CoV-2 molecular testing for asymptomatic screening or diagnosis is performed at Johns Hopkins Diagnostic laboratory by either NeuMoDx (Qiagen) [10, 11], cobas (Roche) [10], Aptima (Hologic), Xpert Xpress SARS-CoV-2/Flu/RSV (Cepheid) [12], ePlex respiratory pathogen panel 2 (Roche) [13], Accula [14], or RealStar SARS-CoV-2 assays (Altona diagnostics) [15]. Testing was performed in accordance with the manufacturer instructions and the Johns Hopkins laboratory’s validated protocols. Patients with greater than 1 positive SARS-CoV-2 test were identified, and were included if final and initial positive tests were >90 days apart. Samples were matched to our sequencing surveillance database for sequencing information.

### Sequencing

Specimens were extracted as previously described [16, 17]. Library preparation was performed with NEBNext ARTIC SARS-CoV-2 companion kit(# E7660-L) with either VarSkip Short (V1 or V2)(New England Biolabs) or Artic V3 primers (Integrated DNA technologies), or as described previously[17]. Oxford Nanopore Technology GridION was used for sequencing, and reads were basecalled with MinKNOW. Demultiplexing was performed with guppy barcoder, with barcoding required at both ends. Alignment, consensus sequence generation and variant calling were performed with Artic using the Medaka pipeline. Mutations requiring further analysis were reviewed with Integrated Genomics Viewer (IGV). Clade determination was performed via NextClade CLI v1.4.5[18].

### Genome analysis

Only high-quality genomes, defined as having a coverage of >90% and depth >100, were used for further analysis. Samples were given an initial determination as reinfection or persistence of initial genome based on whether there was a change in clade assigned by NextClade. Afterward, the consensus genomes for each patient were manually reviewed in NextClade and/or IGV to provide a final determination on reinfection or persistence. The manual review takes into account the initial clade call, the clade call in the later sample, predominance of the clade at the time of the post 90-day positive, presence of any unique mutations, and genome quality as appropriate.

### Statistics

Welch’s t-tests and chi squared analysis were performed to show associations depending on the type of results evaluated. A p-value less than 0.05 was considered significant.

### Study Approval

Research was conducted under Johns Hopkins IRB protocol IRB00221396 with a waiver of consent. Sequencing was performed on remnant clinical specimens from patients that had tested positive for SARS-CoV-2 after clinical testing was performed. Whole genomes were made publicly available at GISAID.

## Results

### Reinfections spiked during December 2021

We Identified 755 patients with two positive SARS-CoV-2 tests at least 90 days apart. Within this cohort of patients, we identified 920 positive tests for SARS-CoV-2 at least 90 days from the initial infection. The second positive test occurred between 91 and 672 days from the initial test, with a median of 377 days, and a peak approximately 1 year from the initial test (Figure 1A). Initial tests in patients who would later have a post 90-day positive were primarily from March-May 2020 period and the winter of 2020/2021, whereas the post 90-day positives were largely collected in late 2021 and early 2022 when Omicron was the dominant variant (Figure 1B). We had attempted sequencing on 231 of the post 90-day positives through our surveillance sequencing efforts, of which 127 whole genomes were recovered from 124 patients. Prior to the Omicron surge, sequencing samples of possible reinfection was only rarely successful. Median time in days from initial positive to post 90-day positive was higher in successfully sequenced samples at 398 days compared to 276 days in samples that failed sequencing (p-value <0.0005, Welch’s t-test) (Figure1A, 1C). While failed samples in the post 90-day positives did increase along with increased testing toward the end of 2021, this was a mild increase compared to the large increase in successfully sequenced isolates post 90-day starting in December of 2021 (Figure 1D).

**Figure 1:**
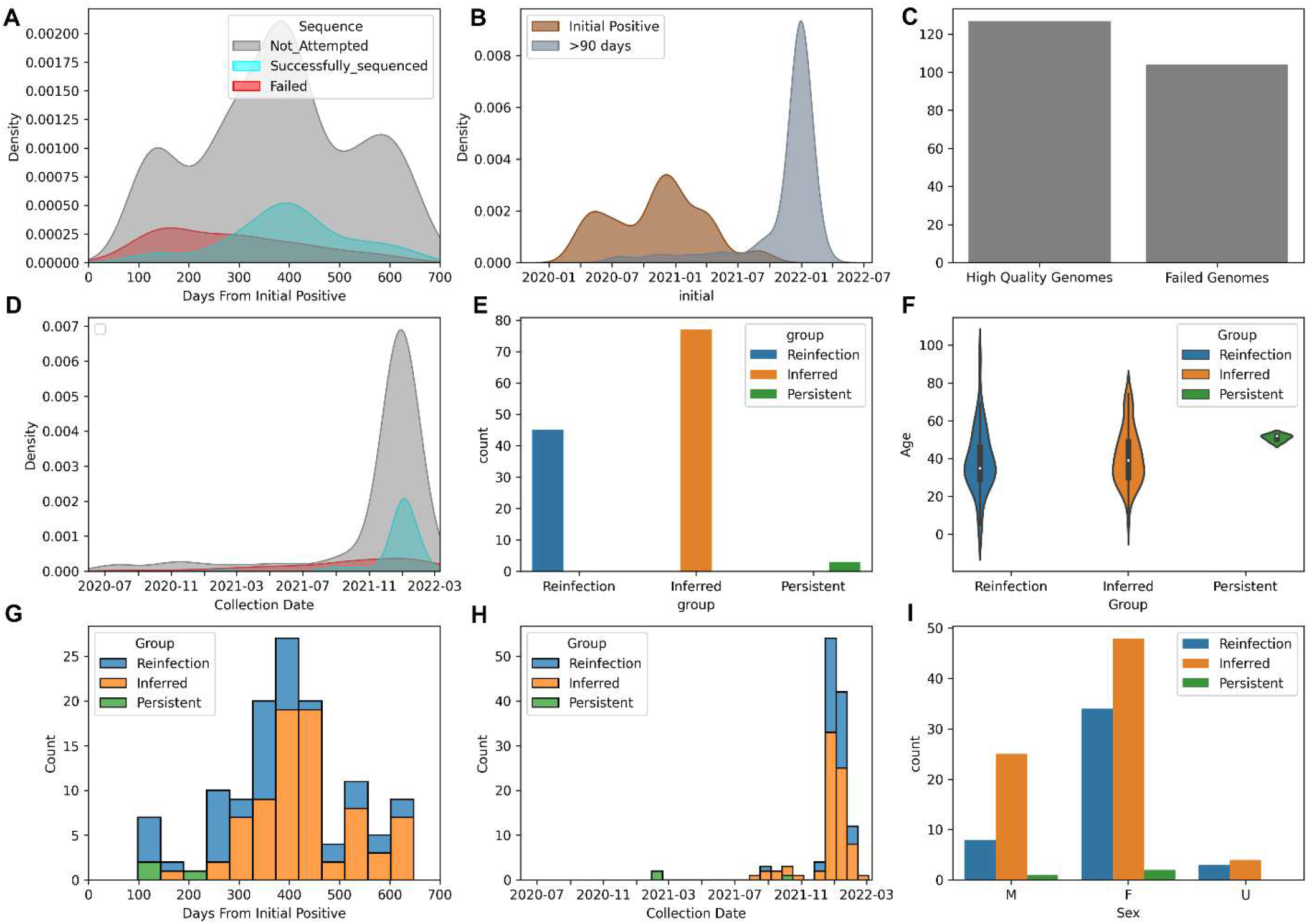
Repeat positive SARS-CoV-2 tests greater than 90 days apart. **A)** Kernel Density Estimator (KDE plot) showing the days from initial infection. Hue indicates whether sequencing was attempted or successful (gray=Not attempted, blue=Pass, Red=fail). (N=920 samples) **B)** KDE plot of date of initial positive and post 90-day positive (N of Post 90-day positive= 920). **C)** Number of tests that failed sequencing or provided high quality genomes (N=231). **D)** KDE plot showing date of sequences that failed or provided high quality genomes(N=920). **E)** Bar plot showing persistence of initial genomes, inferred reinfection, or sequence confirmed reinfection (N=124 patients), **F)** Violin plot showing age and reinfection status in post 90-day positives. **G)** Bar plot showing Days from initial positive to post 90 day positive, hue represents reinfection status (n=127 samples). **H)** Bar plot showing date of post 90 day positive, hue represents reinfection status (n=127 samples). **I)** Bar plot showing sex compared to Reinfection status

Of these patients, 45 had sequence-validated reinfection based on identification of two high quality genomes from different time periods that matched to different Clades (Figure 1E). Additionally, we identified another 77 patients where a high-quality genome in a post 90-day positive sample matched to a clade that did not circulate in this geographical area when the patient was initially infected, and thus we could infer reinfection in these patients. Therefore, within this cohort, >98% of the patients that had a virus that could be sequenced in the post 90-day positive samples, showed evidence of reinfection (122 of 124 patients), compared to 1.6% of genomes (2 of 124 patients) matching to persistence of the original infecting genome.

Patients with post 90-day positive tests who showed persistence of the initial genome showed a range of time from initial sample to post 90-day sample between 111 and 195 days. For sequence proven reinfection, the range was 98 to 646 days with a median of 359 days (Figure 1G). There were no sequence proven or inferred reinfections prior to the emergence of Delta. During the Delta wave, July to November 2021, reinfections were observed on a low, but consistent basis, and then increased several fold during the Omicron surge (Figure 1H). The median age of reinfected patients was 35 years, whereas for patients with persistence this was 52. Children were underrepresented in the reinfection cohort compared to all high-quality Omicron samples in our laboratory (5% compared to 18%, p=0.004 Supplemental Figure 1A). Notably, the failure rate for sequencing was much higher in pediatric patients in the post 90-day positive samples (Supplemental Figure 1B). There were over 4 times as many females with sequence proven reinfection than men in this study (34 females compared to 8 males) (Figure 1I). Females made up approximately twice as many of the post 90-day positives as males (560 compared to 320), (Supplemental Figure 1C), and a higher percentage of samples from females were successfully sequenced compared to those from males, but this was not statistically significant (p=0.19) (Supplemental Figure 1D).

### Reinfections are primarily caused by the Omicron variant and occur in patients who have been vaccinated and are otherwise healthy

Equivalent Delta and Omicron sequences were characterized through our SARS-CoV-2 surveillance initiative (as of the time of writing this manuscript), with Omicron accounting for 29.7% (3282) of total high-quality genomes compared to 26.3% (2914) Delta. Contrastingly, Omicron samples have accounted for 95% of the reinfection cases, and Delta accounted for 5%. (Figure 2A and B). The initial infections in patients who ultimately had reinfections, were caused by all of the major Clades that circulated prior to Omicron. Thirty-two of the patients with sequence proven reinfection were known to be vaccinated prior to the reinfection (Figure 2D), and 12 were known to be unvaccinated. Most patients were symptomatic, whether they were vaccinated or not. The median age was similar between unvaccinated (33 years) and vaccinated patients (36.5 years) (p-value=0.24). The median days between infection and reinfection was also similar between vaccinated and unvaccinated patients (321 days and 368 days respectively). (p-value 0.2). Finally,7 patients had some form of immunosuppression, and the median age of immunosuppressed reinfected patients was higher compared to non-suppressed patients (44 to 34), though this was not statistically significant (p=0.2). Of the 45 patients with sequence proven reinfection, 43 of them were treated in an outpatient setting, and 2 were hospitalized. The two hospitalized patients had underlying conditions contributing to the hospitalization.

**Figure 2:**
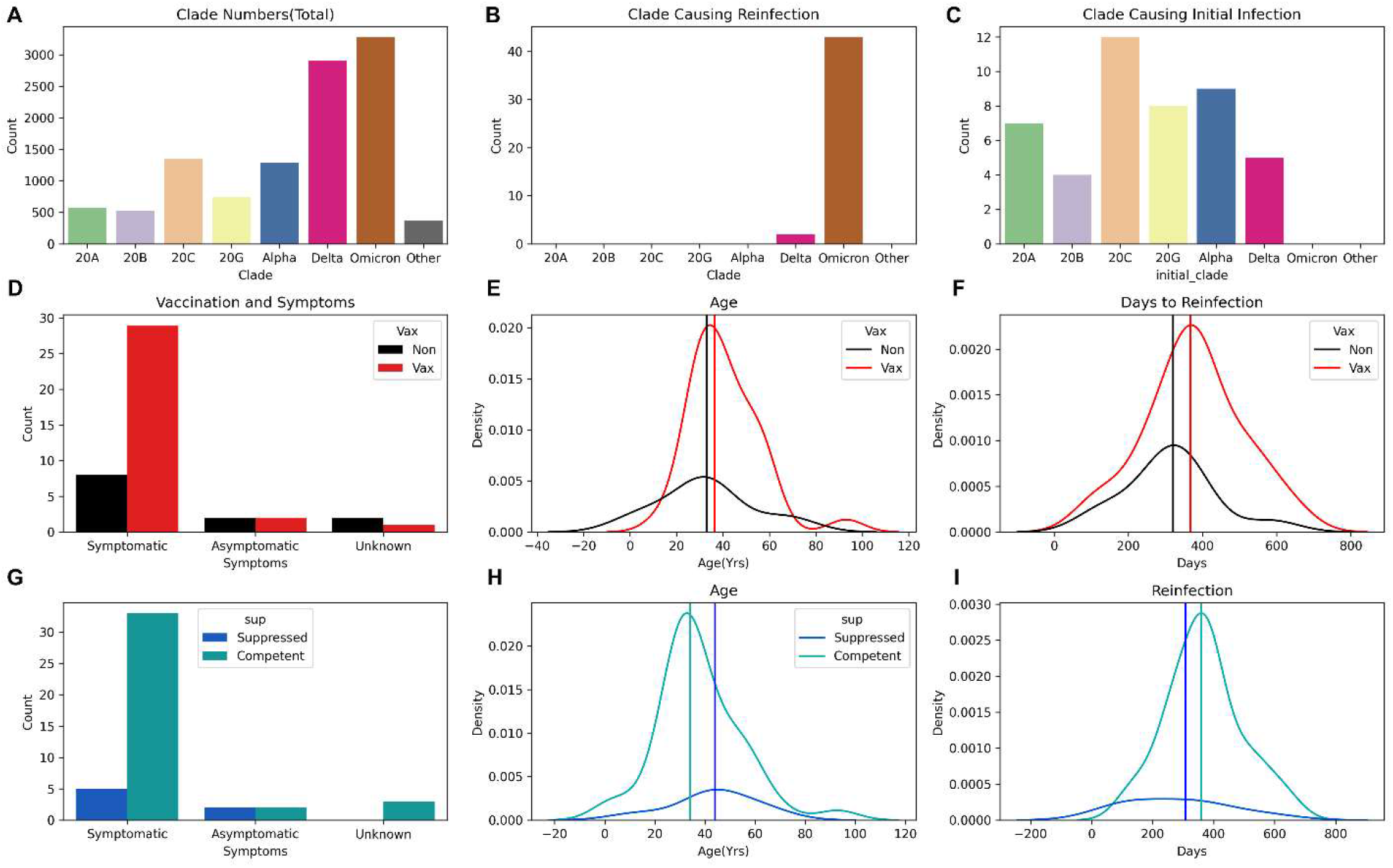
Sequence proven reinfections. **A)** Bar plot of Variant for total high-quality genomes sequenced (n=11024) **B)** Bar plot of variants that have caused sequence proven reinfections. **C)** Bar plot of variants that caused initial infection in sequence proven reinfections. **D)** Bar plot of symptoms in patients with reinfection separated by vaccination status. **E)** KDE plot of age of patients with reinfection separated by vaccine status. **F)** KDE plot of days from initial infection to reinfection by vaccination status. **G)** Bar plot of symptoms in patients with reinfection separated by Immune status. **H)** KDE plot of age of patients with reinfection separated by Immune status. **I)** KDE plot of days from initial infection to reinfection by immune status.

## Discussion

In this study, we showed an increase in the number of sequence proven reinfections starting in late 2021, due mostly to the Omicron variant. Most reinfections occurred approximately a year after the initial infection, and occurred in patients who were vaccinated and immunocompetent. Children were underrepresented among patients with reinfection. As several different groups have noted an increase in the number of reinfections with the Omicron surge based on a second positive test greater than 90 days from initial positive test, we used sequencing data to investigate the patients who met these criteria at the Johns Hopkins System. We noted a large increase in post 90-day positives during the Omicron surge, and sequencing confirmed that >95% of samples that could be sequenced showed evidence of reinfection. Our findings corroborate the high levels of reinfection with the Omicron variant as described by other groups [4], and show that reinfection could occur in vaccinated and immunocompetent individuals. However, sequence proven reinfection rarely led to hospitalization, and was infrequent in children.

There are some limitations to this study. First, there is still a large proportion of genomes that failed sequencing. The reason for this is not always clear, however, successful sequencing is highly dependent on viral RNA load, and thus the failed sequences may be due to lower viral load. However, there is not sufficient evidence to determine the rate at which the failed sequences in the post 90-day positives may be due to reinfections with low viral load or a low-level persistence. It has been shown that residual slowly wanes over approximately a year after an infection [19], which is about the time that sequence proven reinfections spiked after the initial infection. Second, the vast majority of reinfections happened over a small period of time with the Omicron variant. It is clear that the circulating variants have a large impact on the likelihood of reinfection. The Omicron variant is immunologically distinct from prior variants that resulted in incomplete cross protection to Omicron [5]. Thus, as we surveil for reinfections, the likelihood at any given time will likely be dependent on similarity of circulating variants to prior variants. Similarly, although persistence of the initial genome is rare in the post 90-day positives currently, future variants may show a different propensity towards persistence. Continued sequencing is necessary to determine new trends that may arise. Lastly, we cannot obtain a true rate of reinfection compared to total cases with these data, as many patients may have been tested elsewhere for initial infections, or some patients may be reluctant to get tested after having been previously infected.

Interestingly, we show that Immunocompetent, previously infected, young healthy individuals are having reinfections of SARS-CoV-2, primarily with the Omicron variant. Indeed, the majority of samples were patients that were vaccinated and not immunosuppressed, which is consistent with other reports of immune escape by the Omicron variant [20]. The low rates of reinfection in males overall and in children under the age of 18 is somewhat surprising. However, the decreased rates of proven reinfection or inferred reinfection in children may be due simply to low viral load. The evidence for this is that the total number of post 90-day positives was similar in this group compared to adults, but the difference in the ability to prove reinfection was due to an inability to successfully sequence these samples. There remains a large discrepancy between the number of sequence proven reinfections and post 90-day positives in females compared to males, which could represent an increased risk of reinfection for females. However, these findings could also be associated with other variables including the frequency of testing.

Overall, these data suggest that a positive test for SARS-CoV-2 90 days from the initial test was most often due to a repeat infection, and that reinfections can occur in immunocompetent and vaccinated individuals. Despite seeing a large increase in sequence proven reinfection with the Omicron variant, there still appears to be protection from severe disease in this group, as only 2 of the patients with sequence proven reinfections were hospitalized. This is consistent with previous findings from California and New York showing low rates of hospitalization in cases of reinfection [6].

## Data Availability

Genomes are submitted to GISAID.

## Author Contributions

CPM and HHM performed study design. CPM, REE, AF, DCG, JMN, NG, CHL, OA, conducted experiments. CPM, AF, REE, performed data analysis. CPM, HHM, REE, EYK wrote and edited the manuscript. JMN, OA, NG organized samples/experiments. EYK and CPM obtained clinical information. HHM and CPM supervised the study.

## Acknowledgements

This study was only possible with the unique efforts of the Johns Hopkins Clinical Microbiology Laboratory faculty and staff. HHM is supported by the HIV Prevention Trials Network (HPTN) sponsored by the National Institute of Allergy and Infectious Diseases (NIAID). Funding was provided by the Johns Hopkins Center of Excellence in Influenza Research and Surveillance (HHSN272201400007C), National Institute on Drug Abuse, National Institute of Mental Health, and Office of AIDS Research, of the NIH, DHHS (UM1 AI068613), the NIH RADx-Tech program (3U54HL143541-02S2), National Institute of Health RADx-UP initiative (Grant R01 DA045556-04S1), Centers for Disease Control (contract 75D30121C11061), the Johns Hopkins University President’s Fund Research Response, the Johns Hopkins department of Pathology, and the Maryland department of health. EK was supported by Centers for Disease Control and Prevention (CDC) MInD-Healthcare Program (Grant Number U01CK000589). The views expressed in this manuscript are those of the authors and do not necessarily represent the views of the National Institute of Biomedical Imaging and Bioengineering; the National Heart, Lung, and Blood Institute; the National Institutes of Health, or the U.S. Department of Health and Human Services.

---

**Supplemental Figure 1:**
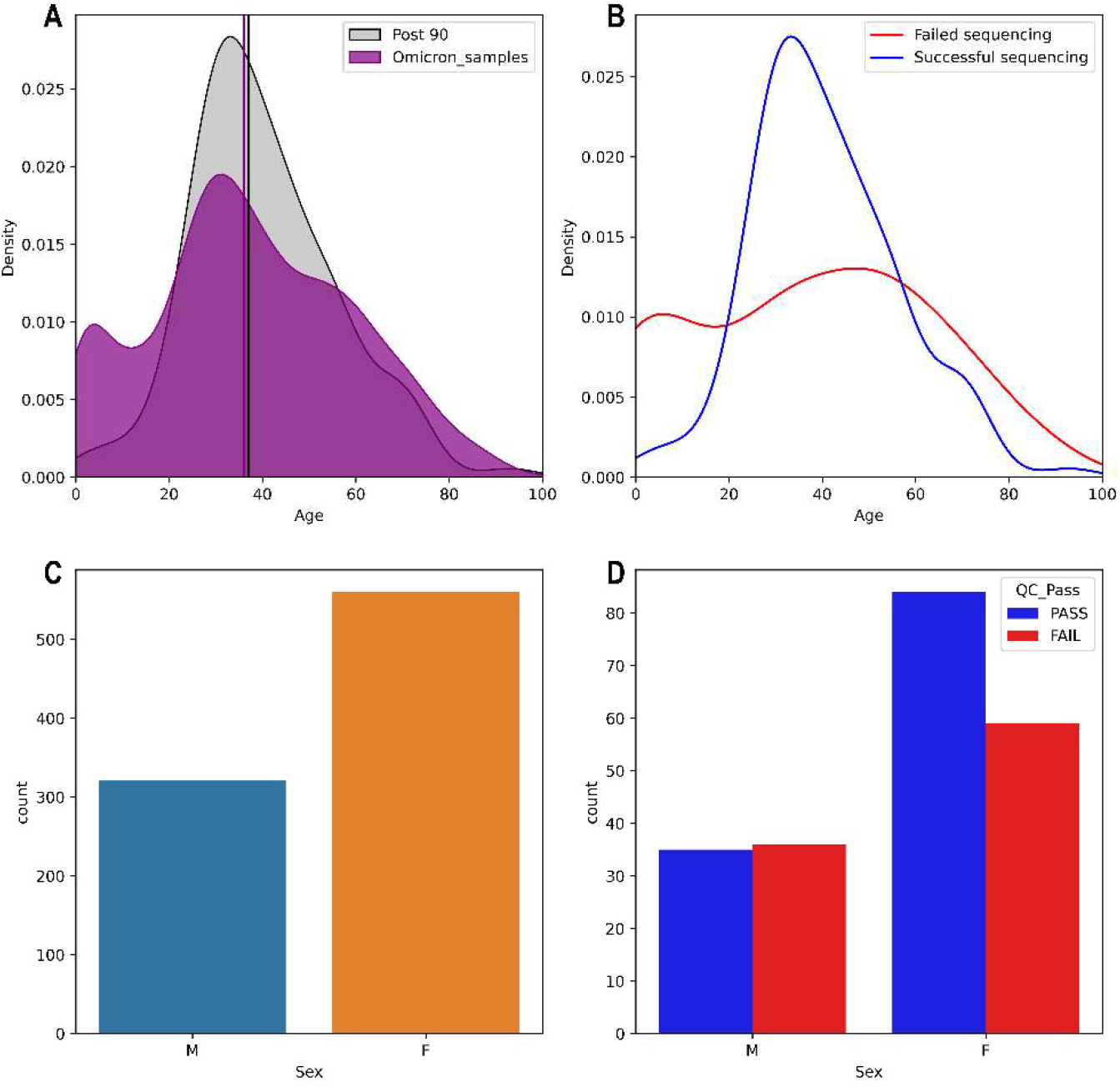
Association of Age and Sex with Successful sequencing, post 90-day positivity, and positivity with Omicron. **A)** KDE plot of age in patients with post 90-day positives (Grey), and Omicron positives (purple). **B)** KDE plot of age of patients in post 90-day positives that failed sequencing (red) and were successfully sequenced (blue). **C)** Count of samples that came from male and female patients in post 90-day positive samples. **D)** Number of post 90-day positives from male and female patients, hue represents whether samples were successfully sequenced (blue) or failed sequencing (red).

## References

1. Fall, A., et al., The displacement of the SARS-CoV-2 variant Delta with Omicron: An investigation of hospital admissions and upper respiratory viral loads. EBioMedicine, 2022. 79: p. 104008.

2. Cele, S., et al., SARS-CoV-2 Omicron has extensive but incomplete escape of Pfizer BNT162b2 elicited neutralization and requires ACE2 for infection. medRxiv, 2021.

3. Khan, K., et al., Omicron infection of vaccinated individuals enhances neutralizing immunity against the Delta variant. medRxiv, 2022.

4. Araf, Y., et al., Omicron variant of SARS-CoV-2: Genomics, transmissibility, and responses to current COVID-19 vaccines. J Med Virol, 2022.

5. Mallapaty, S., COVID reinfections surge during Omicron onslaught. Nature, 2022.

6. León, T.M., et al., COVID-19 Cases and Hospitalizations by COVID-19 Vaccination Status and Previous COVID-19 Diagnosis - California and New York, May-November 2021. MMWR Morb Mortal Wkly Rep, 2022. 71(4): p. 125–131.

7. Long, H., et al., Prolonged viral shedding of SARS-CoV-2 and related factors in symptomatic COVID-19 patients: a prospective study. BMC Infect Dis, 2021. 21(1): p. 1282.

8. Fontana, L.M., et al., Understanding viral shedding of severe acute respiratory coronavirus virus 2 (SARS-CoV-2): Review of current literature. Infect Control Hosp Epidemiol, 2021. 42(6): p. 659–668.

9. Falahi, S. and A. Kenarkoohi, COVID-19 reinfection: prolonged shedding or true reinfection? New Microbes New Infect, 2020. 38: p. 100812.

10. Mostafa, H.H., et al., Comparison of the analytical sensitivity of seven commonly used commercial SARS-CoV-2 automated molecular assays. J Clin Virol, 2020. 130: p. 104578.

11. Mostafa, H.H., et al., Multicenter evaluation of the NeuMoDx™ SARS-CoV-2 Test. J Clin Virol, 2020. 130: p. 104583.

12. Mostafa, H.H., et al., Multicenter Evaluation of the Cepheid Xpert Xpress SARS-CoV-2/Flu/RSV Test. J Clin Microbiol, 2021. 59(3).

13. Jarrett, J., et al., Clinical performance of the GenMark Dx ePlex respiratory pathogen panels for upper and lower respiratory tract infections. J Clin Virol, 2021. 135: p. 104737.

14. Hogan, C.A., et al., Comparison of the Accula SARS-CoV-2 Test with a Laboratory-Developed Assay for Detection of SARS-CoV-2 RNA in Clinical Nasopharyngeal Specimens. J Clin Microbiol, 2020. 58(8).

15. Uhteg, K., et al., Comparing the analytical performance of three SARS-CoV-2 molecular diagnostic assays. J Clin Virol, 2020. 127: p. 104384.

16. Thielen, P.M., et al., Genomic diversity of SARS-CoV-2 during early introduction into the Baltimore-Washington metropolitan area. JCI Insight, 2021. 6(6).

17. Morris, C.P., et al., An Update on Severe Acute Respiratory Syndrome Coronavirus 2 Diversity in the US National Capital Region: Evolution of Novel and Variants of Concern. Clin Infect Dis, 2022. 74(8): p. 1419–1428.

18. Hadfield, J., et al., Nextstrain: real-time tracking of pathogen evolution. Bioinformatics, 2018. 34(23): p. 4121–4123.

19. Pilz, S., et al., SARS-CoV-2 reinfections: Overview of efficacy and duration of natural and hybrid immunity. Environ Res, 2022. 209: p. 112911.

20. Dejnirattisai, W., et al., SARS-CoV-2 Omicron-B.1.1.529 leads to widespread escape from neutralizing antibody responses. Cell, 2022. 185(3): p. 467–484.e15.

